# Characterization of human senescent cell biomarkers for clinical trials

**DOI:** 10.1101/2024.11.02.24316643

**Authors:** Joshua N. Farr, David G. Monroe, Elizabeth J. Atkinson, Mitchell N. Froemming, Ming Ruan, Nathan K. LeBrasseur, Sundeep Khosla

## Abstract

There is an increasing need for blood-based biomarkers of senescent cell burden to facilitate selection of participants for clinical trials. Potential candidates include *p16*^*Ink4a*^ expression in peripheral blood T-cells and circulating protein concentrations of the senescence-associated secretory phenotype (SASP). *p16*^*Ink4a*^ is encoded by the *CDKN2A* locus, which produces six variant transcripts in humans, two of which encode homologous p16 proteins: p16^Inka4a^, encoded by *p16_variant 1*, and p16Ɣ, encoded by *p16_variant 5*. While distinct quantitative polymerase chain reaction primers can be designed for *p16_variant 5*, primers for *p16_variant 1* also measure *p16_variant 5* (*p16_variant 1+5*). In a recent clinical trial evaluating effects of the senolytic combination, dasatinib + quercetin (D+Q), on bone metabolism in postmenopausal women, we found that women in the highest tertile for T-cell expression of *p16_variant 5* had the most robust skeletal responses to D+Q. Importantly, assessment of *p16_variant 5* was more predictive of these responses than *p16_variant 1+5*. Here, we provide a comprehensive *in vitro* and *in vivo* characterization of *p16_variant 5* expression. *In vitro, p16_variant 1* increased rapidly (week 1) following the induction of DNA damage, whereas *p16_variant 5* increased later (week 4), consistent with the latter being more specific for an established senescent state. Further analysis of our clinical trial data identified a SASP panel in plasma that correlated with *p16_variant 5* expression in T-cells and performed as well in identifying postmenopausal women with a positive skeletal response to D+Q. Collectively, our findings support that the assessment of T-cell *p16_variant 5* expression may be more specific for senescence and provide further support for this assay as a biomarker for selecting participants in clinical trials of senolytic interventions. In addition, our data indicate that correlated plasma SASP markers could be used in lieu of the more technically challenging T-cell *p16* assay. Finally, the ability to identify individuals with a beneficial skeletal response to D+Q using two different measures of senescent cell burden (i.e., the T-cell *p16* assay and the SASP score) provides further support for the hypothesis that the underlying senescent cell burden dictates the clinical response to a senolytic intervention.

## Introduction

There is now substantial pre-clinical evidence, principally from mouse models, that clearance of senescent cells ameliorates a range of age-associated co-morbidities [reviewed in Chaib et al.^1^]. This has led to the initiation of a number of early phase clinical trials evaluating senolytic compounds for efficacy in various disease conditions (e.g., Alzheimer’s disease, idiopathic pulmonary fibrosis) as well as otherwise normal aging [reviewed in Raffaele et al.^2^]. A key unresolved issue, however, is how best to select participants in these clinical trials who may have a sufficiently high burden of senescent cells required to respond to senolytic interventions.

This issue was highlighted by a recent clinical trial from our group, which was the first randomized controlled trial (RCT) of a senolytic intervention in humans.^3^ In this phase 2 RCT, 60 postmenopausal women were randomized to intermittent (every four weeks) treatment with the first-generation senolytic combination of dasatinib + quercetin (D+Q) or a control group (n=30 per group) for 20 weeks.^3^ Given our previous preclinical findings that a similar D+Q treatment regimen in old mice reduced bone resorption, increased bone formation, and increased bone mineral density (BMD), along with decreasing senescent cell burden in bone,^4^ our primary and secondary endpoints were changes in the bone resorption marker, C-terminal telopeptide of type 1 collagen (CTx), and in the bone formation marker, procollagen type 1 N-terminal propeptide (P1NP), respectively. Although changes in serum CTx did not differ between groups at 20 weeks, serum P1NP was significantly higher in the D+Q as compared to the control group at 2 and 4 weeks before returning to baseline at 20 weeks – a pattern of changes in bone formation similar to the bone anabolic agent, romosozumab.^5^ Moreover, a key finding of this study that should guide future clinical trials of senolytics was that the skeletal response to D+Q was driven principally by women with a high senescent cell burden where D+Q concomitantly increased P1NP (+34%, *P* = 0.035) and reduced CTx (-11%, *P* = 0.049) at 2 weeks, and increased radius BMD (+2.7%, *P* = 0.004) at 20 weeks.^3^

In that study,^3^ we defined women with a high senescent cell burden as those who were in the highest tertile for peripheral blood T-cell *p16* mRNA levels. This was based on extensive data demonstrating that p16 is a key trigger of cellular senescence^6^ and its expression increases with aging across species.^7-9^ Moreover, studies measuring *p16*^*Ink4a*^ mRNA expression in peripheral blood T-cells not only found an expected age-related increase, but *p16*^*Ink4a*^ expression was also associated with gerontogenic behaviors such as smoking and physical inactivity.^10^ Additional studies found that T-cell *p16*^*Ink4a*^ expression was associated with plasma interleukin-6 (IL-6) levels (a senescence-associated secretory phenotype [SASP] factor),^10^ increased following chemotherapy (along with increases in the SASP factors MCP1 and VEGFA),^11^ and predicted length of hospital stay after coronary artery bypass surgery in older adults.^12^

In establishing our assay for T-cell *p16* mRNA expression, we noted that the human gene encoding p16 (*CDKN2A*) gives rise to six alternatively spliced variants.^13^ The dominant transcript, *p16_variant 1*, produces the established and widely studied p16^Ink4a^ protein. In comparison, *p16_variant 5*, which is expressed at much lower levels, produces the p16Ɣ protein, which is identical to p16^Ink4a^ from amino acids 1-152, but has a unique 15 amino acid C-terminal sequence that replaces the 4 amino acid C-terminal sequence of p16^Ink4a^ (Fig. 1).^3,14^ Fig. 1 also indicates the unique *p16_variant 5* polymerase chain reaction (PCR) primers which measure only the *p16_variant 5* transcript whereas the *p16_variant_1+5* primers measure both *p16_variant 1* and *p16_variant 5* transcripts. Note that the conventional primers used to assess *p16*^*Ink4a*^ mRNA expression in T-cells in fact measure both variant mRNAs.^10^ Importantly, although p16Ɣ is produced at much lower levels than p16^Ink4a^, both proteins appear to have identical functional properties, at least *in vitro*.^14^ Specifically, both proteins interact with CDK4 and inhibit its kinase activity, as well as downstream E2F responses.^14^ The *p16_variants 2, 3, 4, and 6* transcripts either do not encode known proteins or lead to a non-homologous protein (p14^ARF^, encoded by *p16_variant 4*, homologous to p19^ARF^ in mice) with distinct functional properties compared to the two p16 proteins.^13^ Although present in human cells, *p16_variant 5* is not present in mouse cells.^15^

**Figure 1.**
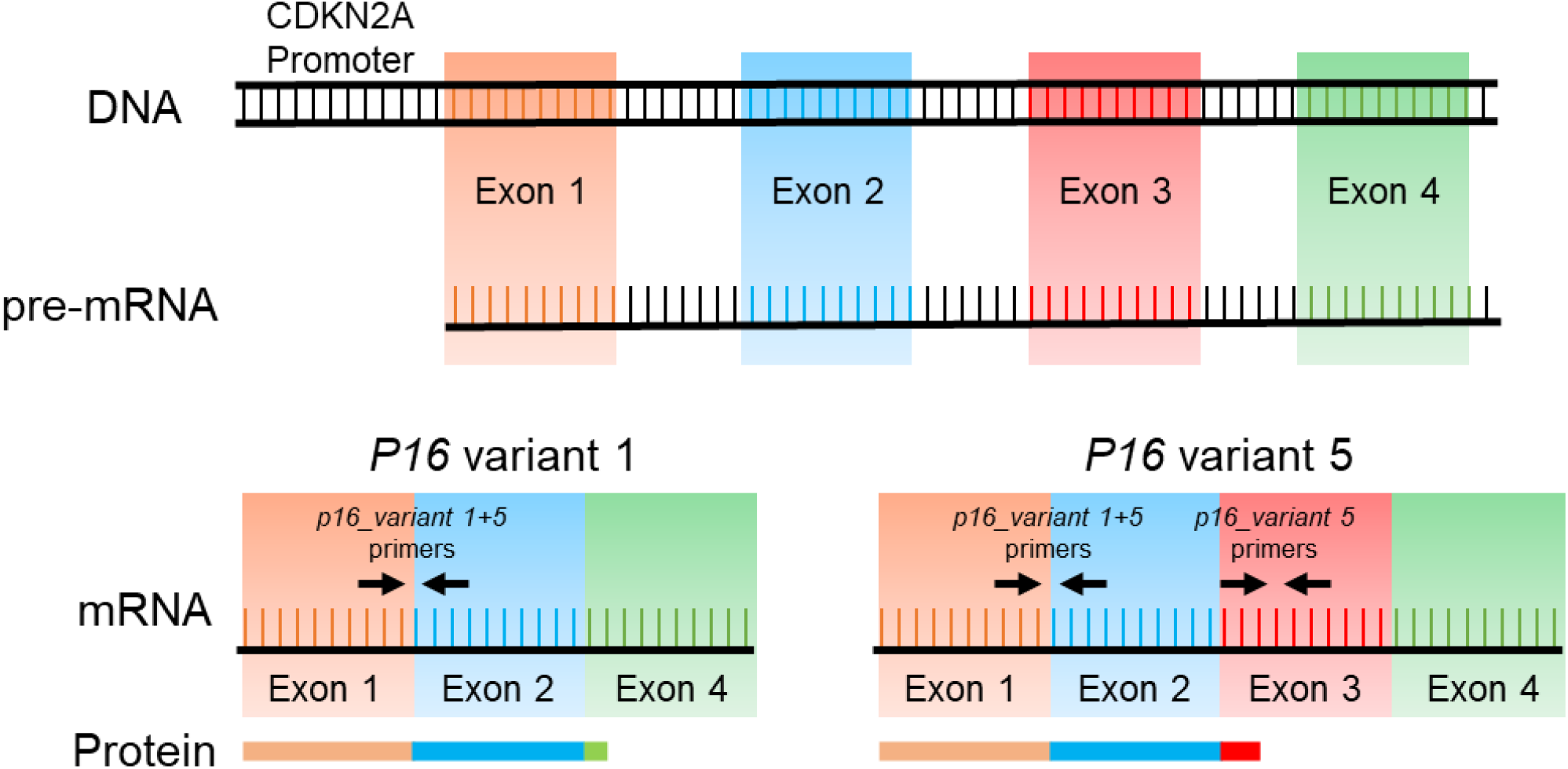
Alternative splicing of the human p16 proximal promoter produces two distinct variants. Schematic representation of the proximal p16 promoter and alternative splicing patterns of the pre-mRNA, indicating the *p16_variant 1* and the *p16_variant 5* mRNAs. The protein products between the two variants encode identical proteins from amino acids 1-152, however the p16_variant 5 protein has a unique 15 amino acid C-terminal sequence in place of the 4 amino acid C-terminal sequence of the p16_variant 1 protein, due to the retained Exon 3 in the *p16_variant 5* mRNA. PCR primer sequences are indicated, demonstrating that the *p16_variant 1 + 5* primer pair amplifies both variant 1 and 5 isoforms, whereas the *p16_variant 5* primer pair only amplifies *p16_variant 5*.

Importantly, in our clinical trial, we found that assessment of *p16_variant 5* was more predictive of skeletal responses to intermittent D+Q treatment as compared to the standard *p16_variant 1+5* assay.^3^ Thus, in the current study, we provide a deeper in vitro and in vivo characterization of *p16_variant 5* expression in human cells and tissues in order to further evaluate the utility of *p16_variant 1+5* versus *p16_variant 5* as potential biomarkers in future clinical trials. Moreover, we reexamine data from our clinical trial^3^ to evaluate whether some combination of plasma SASP factors could substitute for assessing senescent cell burden in lieu of the more technically challenging T-cell *p16* assay.

## Results

### In vitro studies relating induction of p16 variants to senescence

To better understand the expression of *p16* variants following the induction of DNA damage and cellular senescence, we treated human lung fibroblast IMR90 cells with etoposide, a known inducer of DNA damage and senescence.^16^ As shown in Fig. 2A, senescence-associated-β-galactosidase (SA-β-Gal) staining was evident by week 1 following etoposide exposure and was markedly increased at week 4. In order to establish the progression of senescence of these cells, we performed whole-transcriptome RNAseq analyses followed by assessment of the SenMayo gene panel as a validated senescence marker across tissues.^17^ Gene Set Enrichment Analysis (GSEA, Fig. 2B) demonstrated upregulation of the SenMayo gene panel by week 1, with a further increase at week 4 as compared to week 1, consistent with an early DNA damage phase at week 1 progressing to an established senescence phase at week 4. *p21*^*Cip1*^ also increased rapidly (by week 1) and remained elevated at week 4 (Extended Data Fig. 1).

**Figure 2.**
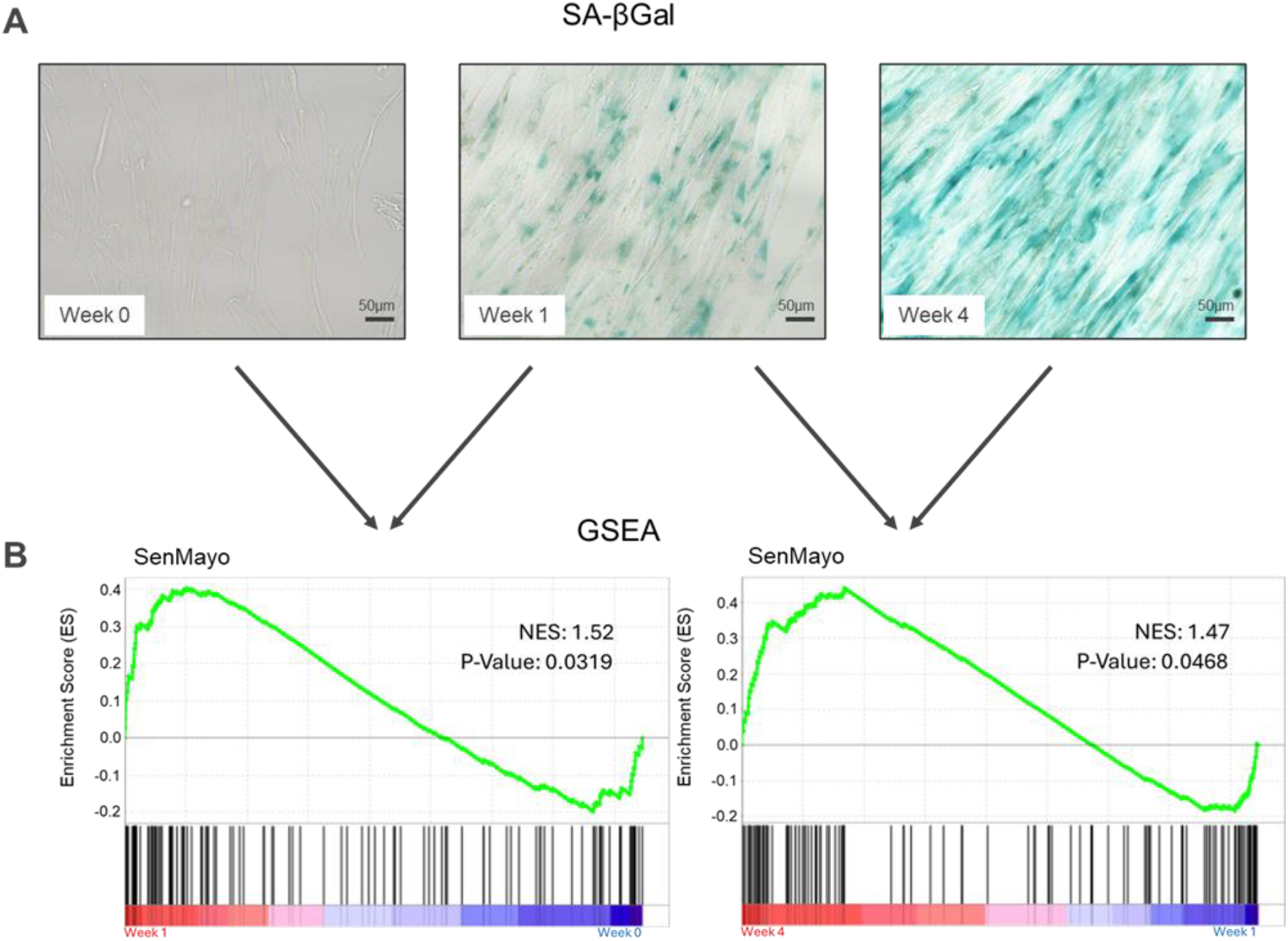
Time course of the DNA damage/senescence phenotype in human IMR 90 cells. (A) Cells were exposed to etoposide and stained for SA-βGal activity 1 and 4 weeks later; (B) RNAseq was performed and the SenMayo gene panel compared using GSEA between the week 0 vs week 1 timepoints and the week 1 vs week 4 timepoints. n=5 week 0, n=6 weeks 1 and 4.

Having defined early DNA damage *versus* established senescence in our in vitro system, we next assessed *p16* mRNA variant expression at these time points. Expression of *p16_variant 1+5* increased rapidly (within a week) of etoposide treatment and remained elevated at week 4 (Fig. 3). By contrast, the *p16_variant 5* mRNA remained unchanged at week 1, but increased significantly at week 4 (Fig. 3). These findings indicate that expression of *p16_variant 1* reflects an acute response to DNA damage that then remains elevated with the establishment of a senescent state, whereas expression of *p16_variant 5* may be more specific for the induction of an established senescent state.

**Figure 3.**
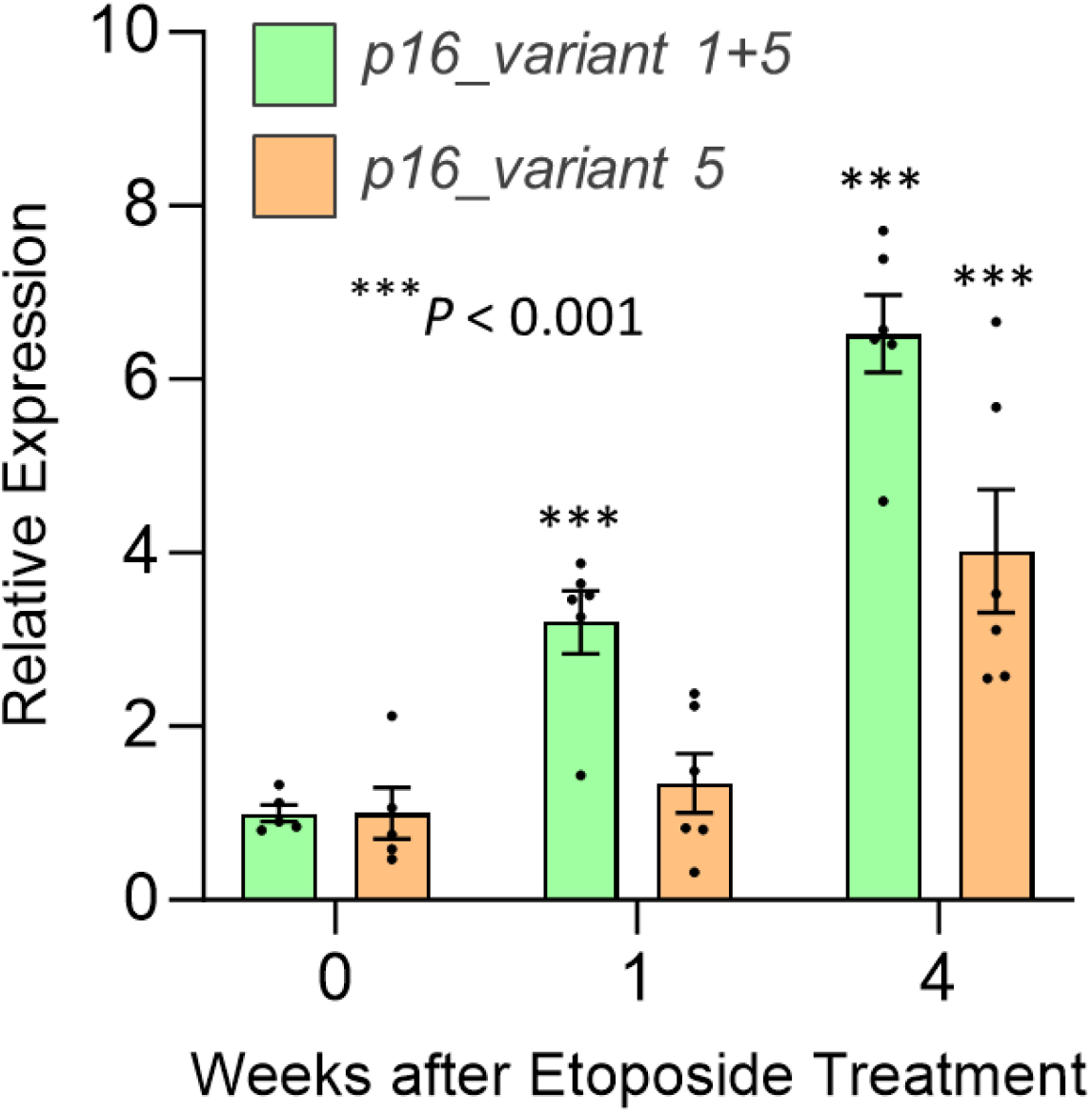
Expression of *p16* variants following *in vitro* induction of DNA damage in human IMR90 cells. Cells were exposed to etoposide to induce DNA damage and expression of *p16_variant 1+5* and *p16_variant 5* mRNAs were assessed using RT-qPCR at baseline (week 0) and weeks 1 and 4. n=5 week 0, n=6 weeks 1 and 4. Data are mean ± SEM; *P-*values based on 2-sided t-test.

### Further evaluation of relationships of age with p16 variants

As noted earlier, in our clinical trial,^3^ the skeletal response to D+Q appeared to be driven principally by women in the highest tertile (T3) for *p16_variant 5* mRNA expression in peripheral blood T-cells. In order to further refine this observation and potentially provide guidance for future trials, we examined the relationships of the *p16_variant 5* tertiles with age. As shown in Fig. 4A, in all of the postmenopausal women aged 62-88 years recruited into the study, only women aged 70 years or older were in the T3 group, whereas all women < 70 years of age were in tertiles 1 or 2 (T1/T2) (Fig. 4B). By contrast, there were a considerable number of women aged 70-88 years who remained in the T1/T2 group (Fig. 4B). Thus, the T3 group consisted exclusively of women 70 years or older, whereas being older than 70 years did not necessarily assure the presence of high levels of *p16_variant 5* mRNA levels in peripheral blood T-cells. Nonetheless, these observations indicate that selecting women ≥ 70 years of age would perhaps be the most efficient way to enrich for a population likely to respond to a senolytic intervention – i.e., women who were in the highest tertile for *p16_variant 5* mRNA expression in T-cells.

**Figure 4.**
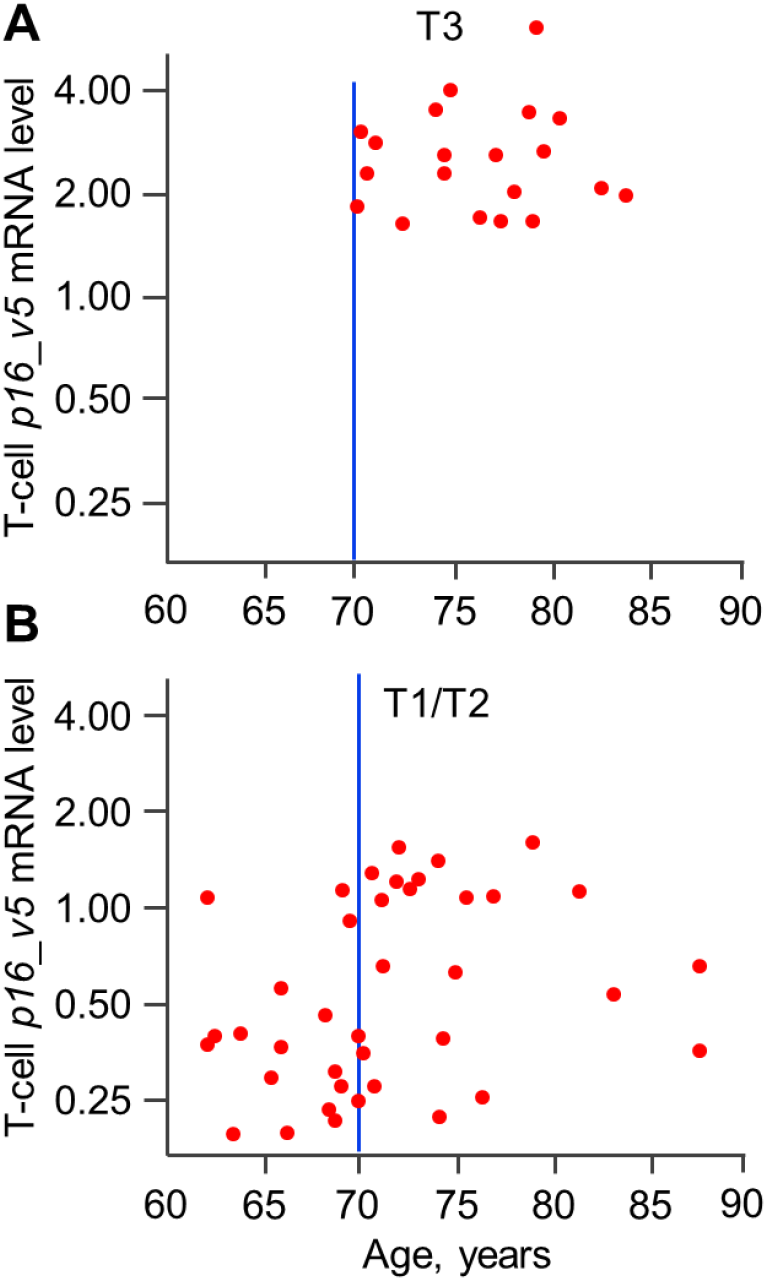
Expression of peripheral blood T-cell *p16_variant 5* mRNA levels as a function of age. (A) Participants in the highest tertile (T3) and (B) participants in the mid- and low-tertiles (T1/T2). Data are adapted from Farr et al.^3^.

### Expression on p16 variants in human bone biopsies

In our previous study,^3^ we found that in peripheral blood T-cells, expression of *p16_variant 5* in women > 60 years correlated more strongly with age (r=0.31, *P=*0.0001) than did expression of *p16_variant 1+5* (r=0.15, *P=*0.044). To extend this to a mesenchymal tissue of interest, we reexamined expression of both *p16* variants in bone biopsies from older women (age 65 to 87 years, n = 20) obtained as part of a previous study.^8^ As shown in Fig. 5, not only in peripheral blood T-cells,^3^ but also in the bone biopsies, expression of *p16_variant 5* correlated much more strongly with age (r=0.46, *P*=0.041; Fig. 5A) than did expression of *p16_variant 1+5* (r=0.27, *P*=0.273; Fig. 5B).

**Figure 5.**
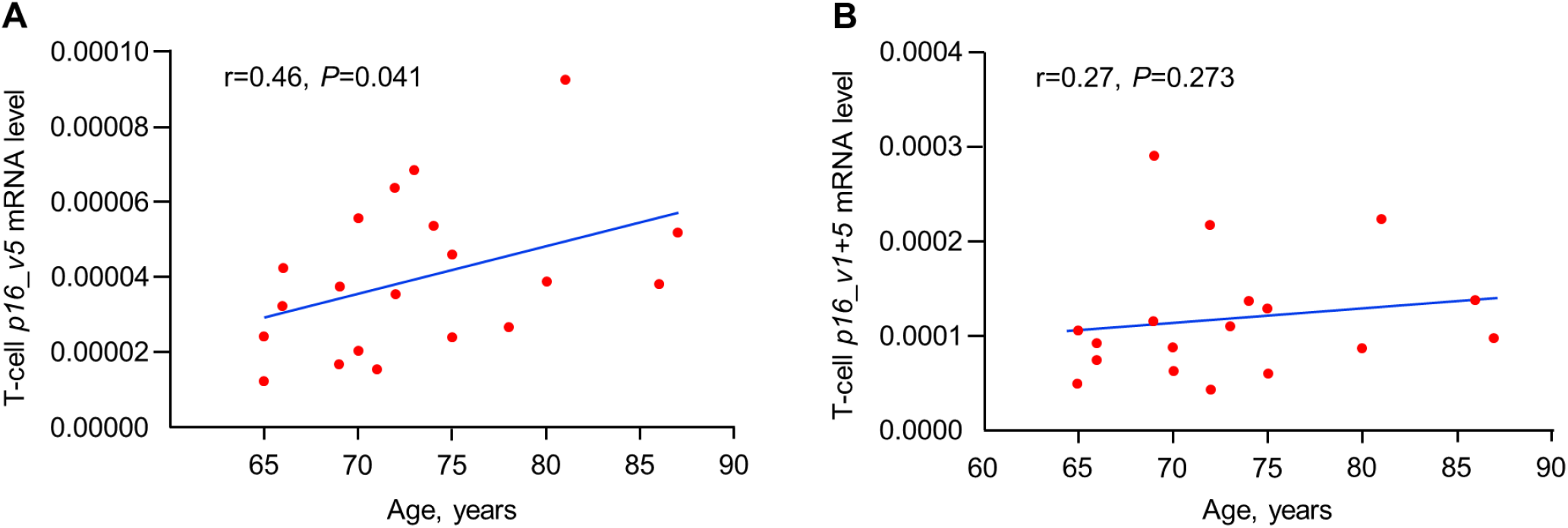
Associations of *p16* variants with age in human bone biopsies. (A) Expression of *p16_variant 5* and (B) *p16_variant 1+5* in relation to age. Correlations are Spearman correlations, n=20.

### Senescence-associated secretory phenotype (SASP) factors as predictors of women in the T3 group

Having established expression of *p16_variant 5* in T-cells as a useful biomarker for assessing senescent cell burden and identifying individuals most likely to respond to a senolytic intervention, we next evaluated whether a subset of plasma SASP factors could be used in lieu of the technically challenging T-cell assay. As part of our clinical trial, we measured a panel of 36 SASP factors at baseline and found that 4 factors (sclerostin, MMP2, Fas, and PARC) were significantly higher in the women in the highest tertile (T3) for T-cell *p16_variant 5* expression as compared to the lower two tertiles (T1/T2); an additional 2 baseline SASP factors (osteoactivin and TNFR1) were higher in the T3 women but were of borderline statistical significance (Extended Data Table 1). We thus created a SASP score for each participant based on the geometric mean of these top 6 SASP factors that differed between the T3 and T1/T2 groups. We then classified the participants into T3 versus T1/T2 groups based on the SASP score rather than the T-cell *p16_variant 5* assay.

As shown in Extended Data Fig. 2, the SASP score was significantly correlated with T-cell *p16_variant 5* mRNA levels (r=0.40, *P=*0.0017). Fig. 6 shows that, similar to classifying participants based on the T-cell *p16_variant 5* assay, classifying them based on the SASP score was also predictive of the T3 group having a robust increase in P1NP at 2-4 weeks following D+Q treatment (Fig. 6A; shaded lines in all Fig. 6 panels show the corresponding changes when using the T3 group for T-cell *p16_variant 5* mRNA levels as done in our previous study^3^), whereas the T1/T2 group failed to respond (Fig. 6B). Of note, the subset of participants who were in the T3 group for both T-cell *p16_variant 5* mRNA levels as well as the SASP score had even more robust increases in serum P1NP at 2-4 weeks (Fig. 6C, D); in fact, the magnitude of differences in P1NP from the control group of ∼60% at these time points is only somewhat less than that observed with the potent sclerostin inhibitor, romosozumab, where these increases were in the 80-90% range.^5^ A similar pattern was observed for CTx (Extended Data Fig. 3A-D) and for radius BMD changes (Extended Data Fig. 4A, B), where the SASP score was generally similar to using the T-cell *p16_variant 5* assay to predict the skeletal response to D+Q, and again the subset of participants who were in the T3 group for both T-cell *p16_variant 5* mRNA levels and the SASP score responded most favorably to senolytic therapy.

**Figure 6.**
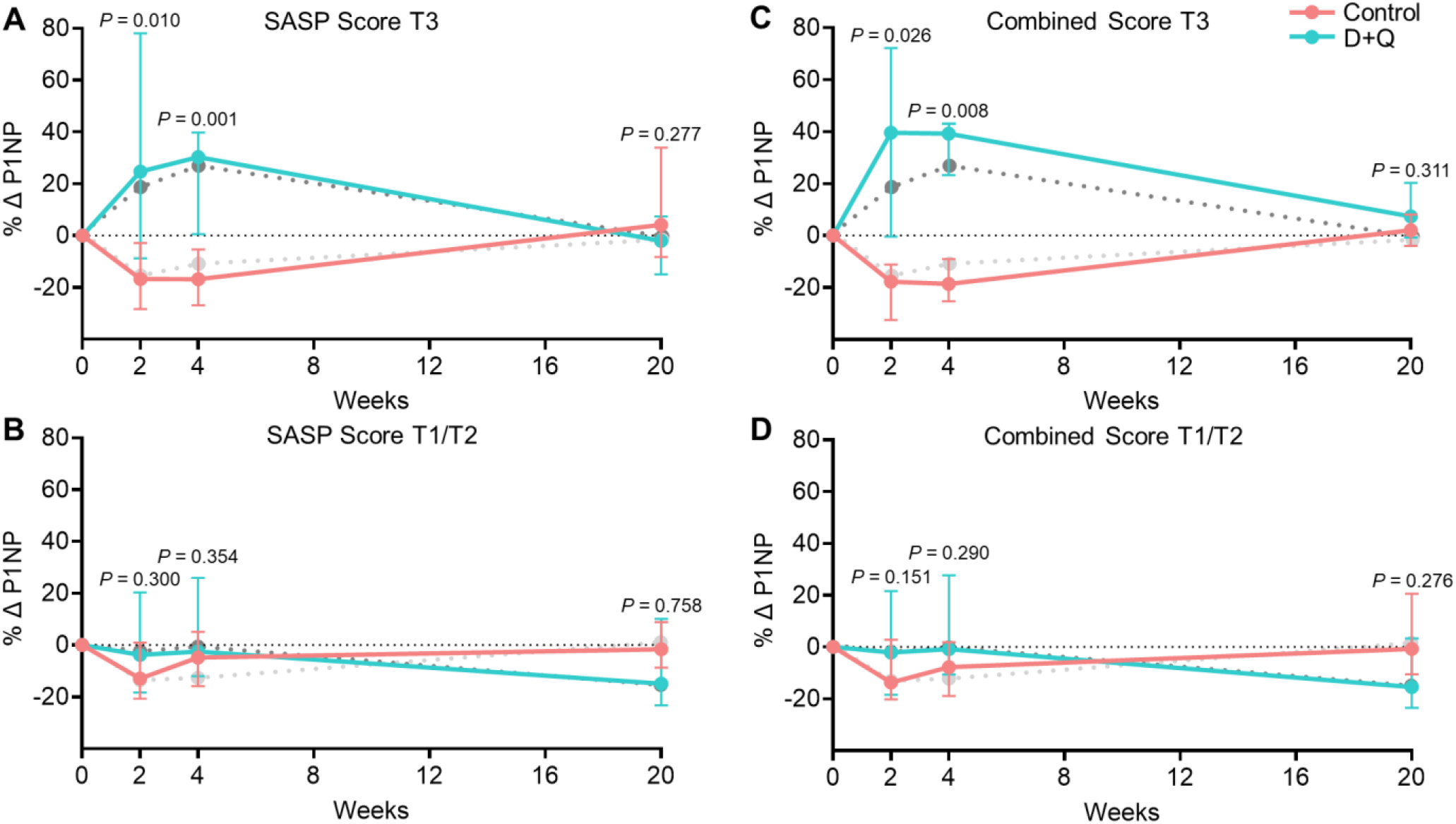
Time course of changes in the bone formation marker, serum P1NP, based on the SASP score tertiles. (A) Participants in the highest tertile for the SASP score (T3); (B) participants in the lower two tertiles for the SASP score (T1/T2), n=21 T3 and 39 T1/T2 at 2 weeks; n=20 T3 and 39 T1/T2 at 4 weeks; n=18 T3 and 38 T1/T2 at 20 weeks; (C) participants in the T3 group for both the SASP score and T-cell *p16_variant 5* mRNA levels; and (D) participants in the T1/T2 groups for either the SASP score and T-cell *p16_variant 5* mRNA levels. n=13 T3 and 47 T1/T2 at 2 weeks; n=12 T3 and 47 T1/T2 at 4 weeks; n=10 T3 and 46 T1/T2 at 20 weeks. In each panel, shaded lines show the corresponding changes based on using the T-cell *p16_variant 5* mRNA for stratification, as per our previous analysis in Farr et al.^3^ (lighter shade is control, darker shade is D+Q). Data are shown as Median (IQR); *P-*values based on two-sided Wilcoxon rank-sum tests.

## Discussion

The present work expands on our previous observation that assessment of peripheral blood T-cell expression of *p16_variant 5* may be more predictive of clinical responses to a senolytic intervention than the standard *p16_variant 1+5* assay.^3^ Our in vitro studies demonstrate that expression of *p16_variant 1+5* increases early following the induction of DNA damage, before the establishment of a senescent state. By contrast, expression of *p16_variant 5* is much more closely linked, at least in human cells (as mice do not express an analogous variant), to an established senescent state. These in vitro data are consistent with our previous observation in a sample of 228 women aged 23-88 years that in younger women (aged 23-60 years) expression of *p16_variant 5* in peripheral blood T-cells was extremely low – when senescent cell burden would be expected to be low, whereas there was substantial spread in the expression of *p16_variant 1+5* in these women.^3^ By contrast, both in our previous study in T-cells^3^ and in the present study in bone biopsies, expression of *p16_variant 5* in women aged > 60 years was more strongly associated with age than expression of *p16_variant 1+5*. Collectively, these in vitro and in vivo observations link expression of *p16_variant 5* much more closely with senescence than expression of *p16_variant 1+5* which, although associated with senescence, may also be responding to other cellular stresses, including early to DNA damage that may or may not cause the cell to progress to a senescent state.

Although the protein products of both *p16_variant 1* (p16^Ink4a^) and *p16_variant 5* (p16Ɣ) appear to have very similar functional properties,^14^ our studies do not address the issue of whether one variant or the other is more effective in inducing a senescent state. Rather, our findings associate established senescence more closely with expression of *p16_variant 5* and provide further support for its utility as a biomarker, not just in T-cells but rather perhaps across human tissues, for assessing senescent cell burden.

Our further analysis of T-cell *p16_variant 5* expression in the study participants from our previous study^3^ also sheds some light on appropriate populations to target for future clinical trials.

Specifically, we demonstrate that all women in the highest tertile (T3) for T-cell *p16_variant 5* expression were ≥ 70 years of age. This is of interest, as using the best estimates to convert human age to mouse age,^18^ age 70 years in humans corresponds to ∼18 months of age in mice – precisely the age at which, in our previous work,^19^ we found that *p16*^*Ink4a*^ expression starts to increase in osteocyte-enriched bone samples from mice. This remarkable concordance between high T-cell *p16* expression in humans and increases in bone *p16* expression in mice further supports that at least in the context of otherwise normal, relatively healthy aging women, targeting participants who were ≥ 70 years of age would enrich for women who were in the T3 group, as these individuals were most likely to have a high senescent cell burden perhaps across tissues, and thus most likely to respond to a senolytic intervention. That said, our estimates indicate that a substantial number (> 50%) of women ≥ 70 years were not in the T3 group (which was originally defined based on age ≥ 60 years^3^), so selecting participants ≥ 70 years would enrich, but not guarantee, participants with high T-cell *p16_variant 5* expression. As such, focusing trials on individuals ≥ 70 years would increase the yield of eligible participants based on further screening for high T-cell *p16_variant 5* expression.

Given the potential logistical obstacles to obtaining T-cell *p16* analyses, including isolating T-cells and performing the qPCR assays, we evaluated whether a SASP score could substitute for the T-cell *p16* assay. Here, using the geometric mean of the top 6 SASP factors that differed at baseline between the T3 and T1/T2 groups by the T-cell *p16_variant 5* assay, we found that the SASP score performed similarly to the T-cell *p16_variant 5* assay in predicting the women who had a beneficial skeletal response to D+Q. Of interest, the sub-group of women who were in the T3 group for both the T-cell *p16_variant 5* assay and the SASP score had the most robust bone formation (P1NP) responses to D+Q, coming close to the P1NP responses observed with the potent sclerostin inhibitor, romozosumab.^5^ It is also important to note that although our endpoints were bone turnover markers, the SASP score defined here was based simply on tertile stratification by baseline levels of circulating protein concentrations of SASP factors, and thus was not intrinsically related to skeletal outcomes. As such, this SASP score may well be generalizable to clinical trials assessing non-skeletal endpoints, although this requires further evaluation.

We have previously addressed possible reasons for the relatively transient increases in P1NP observed following D+Q treatment.^3^ A very similar pattern of an increase and then return to baseline in bone formation markers is observed with other bone-anabolic agents, including romozosumab,^5^ and is likely dictated by the underlying bone biology that is still poorly understood. However, it is also possible that there may be some type of resistance to senolytics that may emerge over time, although in our previous mouse studies, senescent osteocytes remained reduced in bones for up to 4 months following senolytic treatment.^4^

In summary, the present work provides further insights into the potential clinical utility of the peripheral blood T-cell *p16_variant 5* assay for use in selecting and/or stratifying participants in clinical trials. This assay appears to offer greater specificity for assessing senescent cell burden than the standard *p16_variant 1+5* assay, but further studies are needed comparing both assays in population as well as interventional studies. Given the logistical issues in using the T-cell *p16* assay in clinical trials, our SASP score may provide a useful surrogate. Finally, the ability to identify individuals with a beneficial skeletal response to D+Q using two different measures of senescent cell burden (i.e., the T-cell *p16* assay and the SASP score) further supports the hypothesis that the underlying senescent cell burden dictates the clinical response to a senolytic intervention.

## Methods

### Cell culture, etoposide treatments and RNA isolation

IMR-90 human fibroblasts were obtained from the American Type Culture Collection (ATCC; Manassas, VA), cultured in Eagle’s Minimum Essential Medium (EMEM; ATCC) supplemented with 10% (v/v) fetal bovine serum (GeminiBio, West Sacramento, CA), and 1x antibiotic/antimycotic (ThermoFisher Scientific, Waltham, MA) and maintained in a 37^0^C incubator with 5% CO_2_. For the senescence induction experiments, IMR-90 cells were grown to 80% confluence in 6-well dishes and treated with 20 µM etoposide (Sigma-Aldrich, St. Louis, MO) for 48 hours. As a control, untreated cells (Week 0) were collected (n=5) and the etoposide removed from the remaining cells and allowed to grow until harvested for the subsequent timepoints at Weeks 1 and 4 (n=6 each). Total RNA was isolated using the RNeasy Mini Kit (Qiagen, Valencia, CA), which included an on-column DNase step to remove potential contaminating genomic DNA (RNase-Free DNA Set, Qiagen).

### Reverse transcriptase-quantitative polymerase chain reaction (rt-qPCR) analysis

Reverse transcription using 1 µg total RNA per sample was performed using the High-Capacity cDNA Reverse Transcription Kit (Applied Biosystems, Foster City, CA). rt-qPCR was performed on the QuantStudio 7 Real-Time PCR system (ThermoFisher Scientific) using Sybr Green (Qiagen) and primers that recognize human *p16_variant 1* (NM_000077) and *p16_variant 5* (NM_001195132), or primers that specifically recognize *p16_variant 5*. The *ACTB* reference gene was used for normalization. Extended Data Table 2 provides the PCR primers used in this study.

### SA-β-Gal stain

Assessment of *in vitro* senescence was performed using total cellular *SA-β*-Gal activity, performed as previously described ^20^. Briefly, cells either untreated (Week 0) or those treated with etoposide (Week 1 and 4) were fixed in 2% formaldehyde and 25% glutaraldehyde (Sigma-Aldrich) for 5 minutes and were incubated in *SA-β*-Gal staining solution (1 mg/ml X-Gal, 40 mM citric acid, pH 6.0, 5 mM potassium ferrocyanide, 5 mM potassium ferricyanide, 150 mM NaCl, 2 mM MgCl2) for 16 hr at 37^0^C. Following washing in 1x phosphate-buffered saline (pH 7.4), cells were imaged using a EVOS M5000 light microscope at 40 x (ThermoFisher Scientific).

### Bulk mRNA sequencing and analyses

RNA from the etoposide-treated IMR-90 cells harvested at Week 0 (n=5) and Weeks 1 and 4 (n=6 each) from the rt-qPCR analyses were used for whole transcriptome mRNA sequencing at the Mayo Clinic Genome Analysis Core (GAC). Total RNA concentration and quality were determined using Qubit fluorometry (ThermoFisher Scientific, Waltham, MA) and the Agilent Fragment Analyzer (Santa Clara, CA). cDNA libraries were prepared using 100 ng of total RNA according to the manufacturer’s instructions for the Illumina Stranded mRNA Prep, Ligation kit (Illumina, San Diego, CA). The concentration and size distribution of the completed libraries were determined with the Agilent TapeStation’s D1000 ScreenTape and Qubit fluorometry. The cDNA libraries were sequenced at an average read depth of ∼171 million read pairs per sample on an Illumina NovaSeq X Series 1.5B flow cell. Following Illumina’s standard protocol, the flow cell was sequenced with 100 bp x 2 paired end reads using the NovaSeq X Plus Control Software v1.2.2 and RTA4.

The RNA sequencing analysis was performed using the Galaxy open-source web-based platform (https://usegalaxy.org) and includes read trimming, alignment to the hg38 human genome and feature counts to use in downstream analyses. Gene Set Enrichment analysis (GSEA) was performed using the Broad Institute’s GSEA software (GSEA 4.3.2) using feature counts that include only genes with a median expression value ≥ 10 and using run input of 1000 permutations. The SenMayo gene set was used to assess significant alterations in senescence cell signatures in our datasets.^17^

### Study participants

This work did not involve any new study participants, but rather reanalyzed data from a previous studies by Farr et al.^3,8^ Specific information regarding human studies aspects are available there.

### Measurement of T-cell p16 mRNA levels

Data for T-cell *p16_variant 1+5* and *p16_variant 5* analyses in T-cells were obtained from Farr et al.^3^ and from the human bone biopsies from Farr et al.^8^ Detailed methods for sample preparation and RT-qPCR analyses are provided in those publications.

### Measurement of serum SASP proteins

SASP proteins were measured in patient morning fasting plasma samples obtained at baseline and at 2 weeks using commercially available multiplex magnetic bead immunoassays (R&D Systems) based on a Luminex xMAP multianalyte profiling platform, as described in Farr et al.^3^

### Statistical analyses

Unless otherwise specified, all data were presented as median (interquartile range). The clinical trial study endpoints were expressed as a percentage of baseline for the Figures and statistical analyses. Comparisons between the control and D+Q groups were made using the Wilcoxon rank-sum test. Spearman correlation coefficients were used to summarize relationships between continuous variables. For the in vitro studies, data are indicated as mean ± SEM, and 2-sided t-tests were used. A P-value of <0.05 was considered statistically significant.

### Statistics and reproducibility

Replicates for the in vitro studies are noted in the manuscript. In the reanalysis of the human trial data, the experiments were conducted once and all relevant data are presented.

### Reporting summary

Further information on research design is available in the Nature Portfolio Reporting Summary linked to this article.

## Data Availability

All information on materials and reagents is provided in the Methods. Individual deidentified participant data that underlie the results reported in this article (text, tables, figures) will be made available as an Excel file in the Supplementary Information upon acceptance. The study used reanalysis of previous clinical trial data from Farr et al.(PMID: 38956196), so the study protocol and IRB information are provided in that publication.

## Data availability

All information on materials and reagents is provided in the Methods. Individual deidentified participant data that underlie the results reported in this article (text, tables, figures) will be made available as an Excel file in the Supplementary Information upon acceptance. The study used reanalysis of previous clinical trial data from Farr et al.,^3^ so the study protocol and IRB information are provided in that publication.

## Code Availability

Not applicable – no code used.

## Acknowledgements

This work was supported by National Institutes of Health grants R21 AG065868 (J.N.F. and S.K.), P01 AG062413 (S.K., D.G.M.), R01 AG076515 (S.K., D.G.M.), R01 AG086085 (S.K., D.G.M.), Hevolution HR-GRO-23-1199144-8 (S.K., D.G.M.), and R01 DK128552 (J.N.F.). The funders played no role in the conduct or analyses of this study.

## Author contributions

J.N.F., D.G.M. and S.K. conceived and directed the project. D.G.M., M.N.F., and M.R. conducted the experimental analyses. N.K.L.’s laboratory performed the SASP analyses. J.N.F., D.G.M., M.N.F., and E.J.A. performed the statistical analyses. S.K. and J.N.F. wrote the manuscript, which all authors then reviewed and approved.

## Conflict of interest statement

The authors have no conflicts to declare.

### Ethical Statement

The study used reanalysis of previous clinical trial data from Farr et al.,^3^ so the study protocol and IRB information are provided in that publication

