## Extended Data for "Characterization of human senescent cell biomarkers for clinical trials"

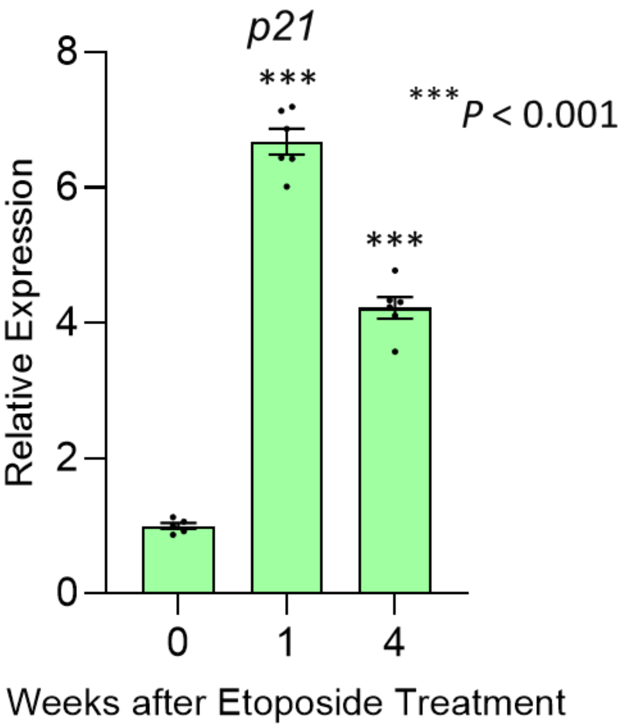


**Extended Data Figure 1. Changes in *p21$^{Cip1}$* mRNA levels in IMR90 cells.** Cells were exposed to etoposide to induce DNA damage and expression of *p21$^{Cip1}$* mRNA levels were assessed using RT-qPCR at baseline (week 0) and weeks 1 and 4. n=5 week 0, n=6 weeks 1 and 4. Data are mean ± SEM; *P*-values based on 2-sided t-test.

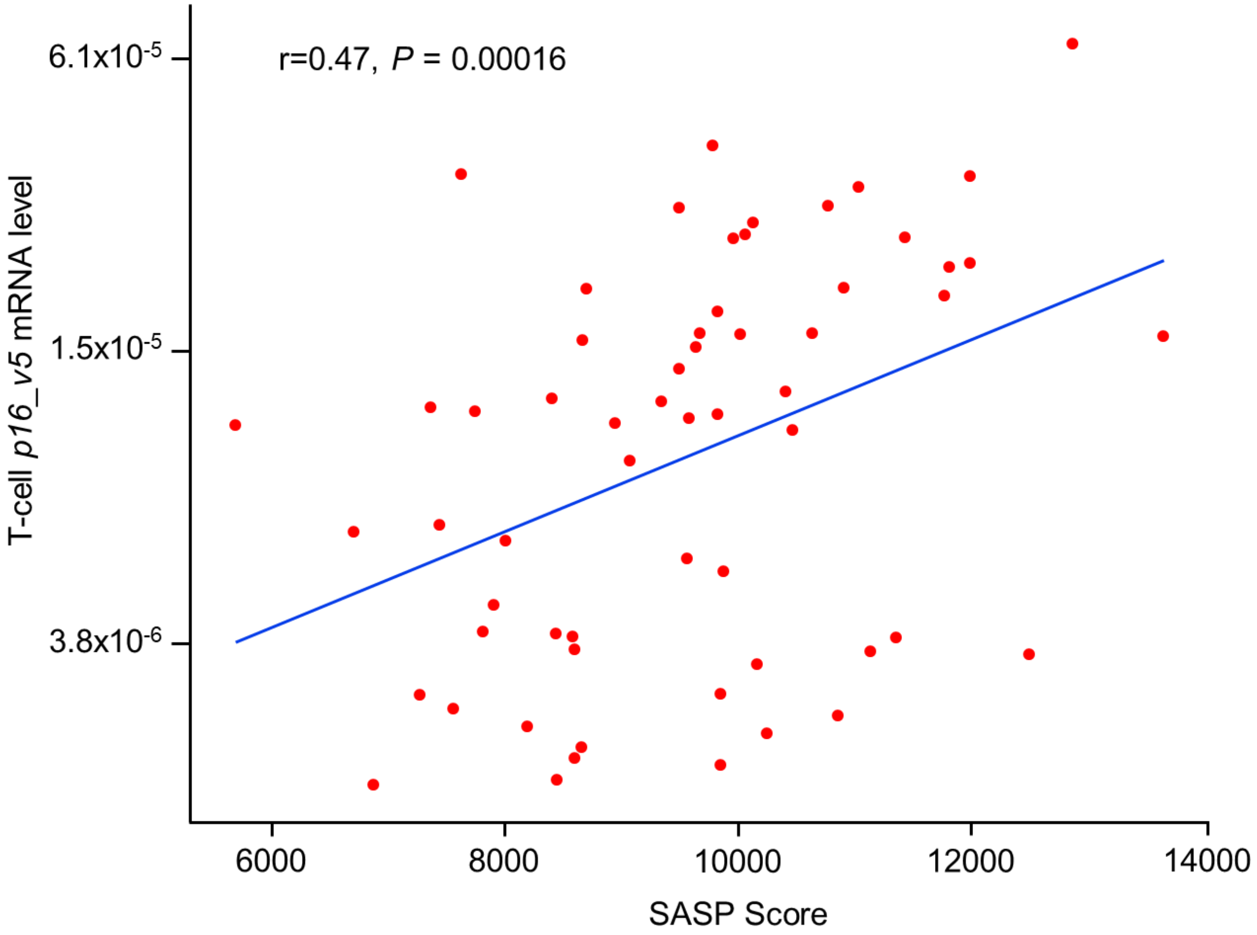


**Extended Data Figure 2. Correlation between the SASP score and T-cell *p16_variant 5* mRNA levels.** N=60, Spearman correlations.

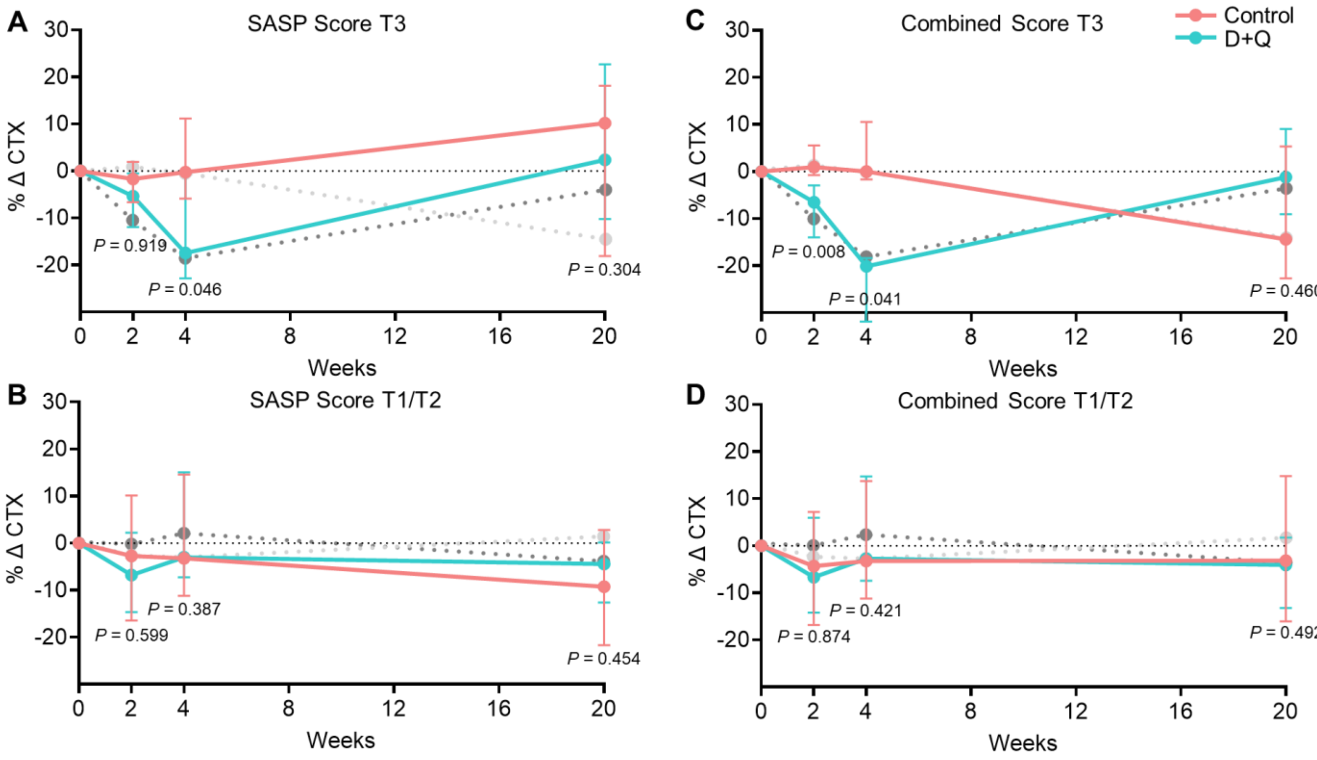


**Extended Data Figure 3. Time course of changes in the bone resorption marker, serum CTx, based on the SASP score tertiles.** (A) Participants in the highest tertile for the SASP score (T3); (B) participants in the lower two tertiles for the SASP score (T1/T2), n=21 T3 and 39 T1/T2 at 2 weeks; n=20 T3 and 39 T1/T2 at 4 weeks; n=18 T3 and 38 T1/T2 at 20 weeks; (C) participants in the T3 group for both the SASP score and T-cell *p16_variant 5* mRNA levels; and (D) participants in the T1/T2 groups for either the SASP score and T-cell *p16_variant 5* mRNA levels. n=13 T3 and 47 T1/T2 at 2 weeks; n=12 T3 and 47 T1/T2 at 4 weeks; n=10 T3 and 46 T1/T2 at 20 weeks. In each panel, shaded lines show the corresponding changes based on using the T-cell *p16_variant 5* mRNA for stratification, as per our previous analysis in Farr et al.[3] (lighter shade is control, darker shade is D+Q). Data are shown as Median (IQR); *P*-values based on two-sided Wilcoxon rank-sum tests.

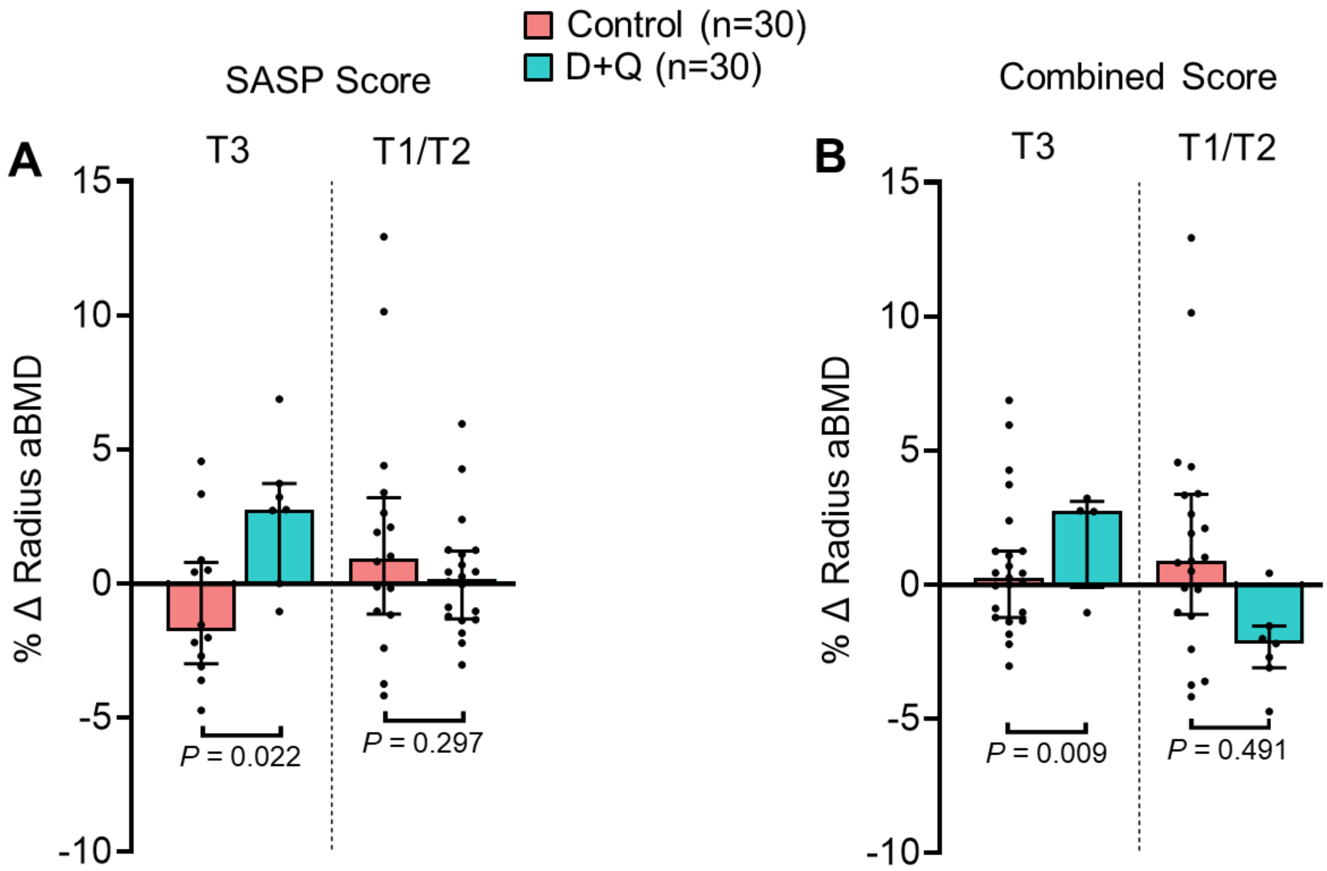


**Extended Data Figure 4. Changes in radius BMD at 20 weeks based on the SASP score tertiles.** (A) Participants in the highest tertile for the SASP score (T3) versus participants in the lower two tertiles for the SASP score (T1/T2), n=19 T3 and 38 T1/T2; (B) participants in the T3 group for both the SASP score and T-cell *p16_variant 5* mRNA levels versus participants in the T1/T2 groups for either the SASP score and T-cell *p16_variant 5* mRNA levels, n=11 T3 and 44 T1/T2. Data are shown as Median (IQR); *P*-values based on two-sided Wilcoxon rank-sum tests.

**Extended Data Table 1. Top 6 circulating SASP factors in the study subjects from Farr et al.[3] that differed between the T1/T2 and T3 groups.**

| | **T1/T2** | **T3** | ***P*-value** |
|---|---|---|---|
| Sclerostin, pg/mL | 721 (629, 862) | 960 (869, 1137) | 0.005 |
| MMP2, pg/mL | 110579 (87604, 128699) | 127805 (109472, 134052) | 0.017 |
| Fas, pg/mL | 6790 (5800, 7815) | 7444 (6875, 8900) | 0.034 |
| PARC, pg/mL | 43085 (30408, 61976) | 54721 (44366, 67142) | 0.045 |
| Osteoactivin, pg/mL | 18764 (15856, 20836) | 20467 (18481, 22421) | 0.056 |
| TNFR1, pg/mL | 1216 (1001, 1359) | 1345 (1206, 1506) | 0.064 |

Results are shown as Median (IQR); *P*-values based on two-sided Wilcoxon rank-sum tests.

**Extended Data Table 2. qPCR primer sequences used in the study.**

| Gene | Description | Forward Primer Sequence | Reverse Primer Sequence |
|---|---|---|---|
| *ACTB* | Housekeeping | CCCAGCCATGTACGTTGCTAT | TCACCGGAGTCCATCACGAT |
| *CDKN2A_variant 5* | *p16_variant 5* | CAGAAATGATCGGAAACCATT | CTACGCATGCCTGCTTCTAC |
| *CDKN2A_variant 1+5* | *p16_variant 1+5* | CCAACGCACCGAATAGTTACG | GCGCTGCCCATCATCATG |
